# The link between CS gas exposure and menstrual cycle issues among female Yellow Vest protesters in France

**DOI:** 10.1101/2020.10.11.20210955

**Authors:** Yara Mahfud, Alexander Samuel, Elif Çelebi, Jais Adam-Troian

## Abstract

CS teargas is one of the most used tools for crowd-control worldwide. Exposure to CS teargas is known to have consequences on protesters’ health (i.e. eye, skin irritation, respiratory problems), but recent concerns have been raised over its potential gender-specific effects. Indeed, field and clinical observations report cases of menstrual cycle issues among female protesters following high exposure to teargas. The hypothesis of a link between teargas exposure and menstrual cycle issues is plausible from a physiological standpoint, but has not yet been empirically investigated. Using data from a cross-sectional study on Yellow Vests protesters’ health in France, we examined the relationship between exposure to teargas and menstrual cycle issues among female protesters (*n* = 145). Analyses suggested a positive link between exposure and menstrual cycle perturbations. These results constitute first and preliminary evidence that CS teargas may be linked with menstrual cycle among women, which need corroboration given the importance of this issue. We call for further research on the potential effects of CS teargas on women’s reproductive system.

## 1. INTRODUCTION

A surge in mass protests has been observed throughout the world over the past two years. From the Yellow Vest in France (Jetten, Mols, & Selvanathan, 2020) to the Hong-Kong anti-extradition bill and the more recent ‘George Floyd’ protests in the US, a common thread has been the level of police response deployed by governments. More specifically, these responses made use of crowd control techniques relying on various weapons, which can have tremendous effects on protesters’ health. For instance, kinetic weapons (i.e. rubber pellet ammunition launchers) were found to cause severe ocular injuries, sometimes leading to permanent loss of eyesight (Chauvin et al., 2019). Stun grenades and other explosive devices used to disperse crowds have caused mutilation and loss of hands among protesters (Jetten et al., 2020; Paye, 2019). Likewise, it has recently been acknowledged that police use of force and less-lethal weapons could have deleterious effects on participants’ mental health (Celebi, Adam-Troian, & Mahfud, 2020)

Among the different tools available to law enforcement the most widely used remains Ortho-Chlorobenzylidene Malononitrile, also simply known as CS teargas or teargas. As an example, more than 16,000 rounds of teargas were fired during the 2019 Hong Kong protests (Standard, 2019). In Paris, France, 7940 teargas grenades have been used in a single day of December 1^st^ 2018 during the Yellow Vest protests (Rocher, 2020). Despite its widespread use by law enforcement, tear gas comprises many known health hazards, with scientists calling for restriction of its use (Haar, Iacopino, Ranadive, Weiser, & Dandu, 2017; Kaszeta, 2019; Rothenberg, Achanta, Svendsen, & Jordt, 2016). Cutaneous effects vary from transient rash and erythema to severe allergic dermatitis (Varma & Holt, 2001) with second to third degree burns (Zekri, King, Yeung, & Taylor, 1995). Eye damage includes corneal edema, keratitis, glaucoma and even cataract (Gray & Murray, 1995; Hout, White, Stubner, Stevens, & Knapik, 2014). On the respiratory tract, effects vary from cough to airway obstruction with hemoptysis (Dagli, Uslu, Ozkan, Torlak, & Arbak, 2014) and damage induces higher susceptibility to acute respiratory inflammations (Hout et al., 2014), which is of high concern during COVID-19 pandemic.

Besides these general consequences of teargas on the organism, there are reasons to suspect that gender specific effects may occur - which remain understudied. More specifically, recent concerns have been raised about teargas’ potential effect on the female menstrual cycle. Reports from recent press releases indicate that these concerns are more than simple public rumor (Anizon, 2019) and a survey study run by Planned Parenthood North Central States and the University of Minnesota was started this year (Hassan, 2020) to assess whether teargas may affect female protesters’ menstrual cycle in particular (longer or shorter cycles). Indeed, clinical observations of increases in miscarriages and stillbirths after tear gas exposure have been reported in Gaza (Hu et al., 1989) and in Bahrain (Atkinson & Sollom, 2012). Concerns over potential miscarriages have led some countries to suspend the use of teargas, like Chile in 2011 (Rothenberg et al., 2016; also see Hayman, 2011).

Still, the current level of evidence for a link between CS teargas and menstrual/reproductive perturbations is low, although the hypothesis remains physiological plausible. In fact, a pathway leading to these symptoms based on CS metabolism into cyanide (Frankenberg & Sorbo, 1973) after absorption (Leadbeater, 1973) or direct cyanide exposure after thermal degradation (Johnson-Kanapathy, 2013) was recently proposed (Samuel et al., 2020). Although this hypothesis has not yet been tested, teargas could affect women’s menstrual cycle through a molecular dysfunction due to decreased blood level oxygen (a consequence of increased cyanide blood levels). This would be consistent with studies showing that hypoxia leads to impaired menstrual cycle among humans (Maybin et al., 2018).

In addition, injection of CS in rodents was found to increase blood thiocyanate (the final product of cyanide in the metabolism (Ballantyne, 1983). Thiocyanate being a perturbator of the thyroid function (Dohan, De la Vieja, & Carrasco, 2000), and thyroid dysfunction being related to menstrual disorders (Ajmani et al., 2016), this could be another pathway through which teargas may affect women’s menstrual cycle. Converging evidence also suggests that other teargas affected molecular mechanisms could trigger menstrual cycle impairment, either due to byproducts or to a potential endocrine disruptive effect of teargas (Nelson, Wang, Sakwari, & Ding, 2020). In sum, various physiological mechanisms triggered by teargas could-theoretically - act or interact to produce perturbations on the female menstrual cycle.

Despite growing concerns, mounting clinical evidence and biological plausibility, the potential effect of CS teargas on female menstrual cycle has - to the best of our knowledge - not been investigated yet. We therefore decided to test whether teargas exposure could be associated with menstrual cycle perturbations among female protesters. To do so, we took advantage of data collected from a study on protester health and social behavior just before the beginning of the covid-19 crisis. We hypothesized that levels of teargas exposure among female protesters would be positively associated with prevalence of menstrual cycle perturbation symptoms.

## 2. METHODS

### 2.1. Ethical statement

The study was conducted in accordance with the 1964 Helsinki declaration and its later amendments, the French legislation on research involving human participants and the 2016 APA Ethical Principles of Psychologists and Code of Conduct. Ethic approval was obtained from [ANONYMIZED INSTITUTION] ethics board (n°29-2019). The data underlying our findings can be openly accessed using the following link https://osf.io/k3upf/?view_only=39158ca00ff84a03ad9d3b41691abbba

### 2.2. Participants and procedure

Our data comes from a larger cross-sectional study assessing the various effects of crowd control techniques on protesters’ health and behavior in France. In this context, we aimed to collect data from a sample of individuals involved in the Yellow Vests protests, albeit limited to those individuals able to access online information and to participate in online surveys. To ensure stable parameter estimates, we estimated that at least 250 respondents were needed (Schönbrodt & Perugini, 2013).

Links to an online survey were disseminated on Yellow Vests social media pages by the investigators. Leaders of the movement and administrators of webpages and forums dedicated to the Yellow Vests were also asked to voluntarily help disseminate survey links. Data collection started in February of 2020. Investigators frequently monitored the sample size and were instructed to stop collection when the expected sample size of at least 250 Yellow Vest participants was reached. In march of 2020, the COVID-19 pandemic started in France and data collection was thus interrupted to avoid bias. Therefore, the survey contained responses from 215 self-identified Yellow Vests (67.8% female; *M*_age_ = 47.6, *SD* = 11.2), who attended 26.3 Yellow Vests protests on average (*SD* = 26.2). Among them, our final sample for the current study is made of the n = 145 women who responded to the survey (*M*_age_ = 48.6, *SD* = 10.2), who attended 23.8 protests on average (*SD* = 23.4).

### 2.3 Materials

The survey contained two parts, one epidemiological and one psychosocial, for different studies and investigative purposes. The psychosocial part of the survey included measures related to political attitudes and protest behavior. The epidemiological part of the survey examined in this study contained measures of health, medical antecedents, mental health as well as a specific part related to exposure to teargas and teargas-related symptoms. Among them, we used the following measures:

#### Exposure to teargas

A single item measure was used to assess the extent to which participants had been exposed to teargas. The item asked how often participants had been exposed to teargas in the protests (0 = ‘*Never*’, 1 = ‘*Less than 5 times*’, 2 = ‘*Between five to ten times*’, 3 = ‘*More than 10 times*’; *Median* = 2). Overall, 19.6% (n = 28) of the participants were never exposed to teargas, 21% (n = 30) less than five times, 14.7% (n = 21) between five and ten times and 44.8% (n = 64) more than ten times.

#### Teargas-specific symptoms

Irritating effects, achieved through transient receptor potential ankyrin 1 (TRPA1) receptors stimulation (Lindsay et al., 2015) are mainly on the skin, eyes and respiratory tract (Schep, Slaughter, & McBride, 2015). Cyanide effects are those of hypoxia, causing headaches, dizziness, confusion, coma or even death (L. Nelson, 2006). Oxidative stress caused by cyanide poisoning can turn into long lasting effects on liver, kidneys and brain (Bhattacharya, Singh, John, & Gujar, 2018). These teargas specific symptoms (ranging from eye burn to nausea and heart problems; n = 15) were measured by asking participants if they experienced any of those following exposure to teargas. Furthermore, participants were asked to indicate if they experienced these symptoms directly after exposure (short term) or if these still persisted a few days after exposure (long term). From these, we computed scores of total reported (*M* = 7.13, *SD* = 5.18) and long-term only symptoms (*M* = 2.92, *SD* = 3.56) following teargas exposure.

#### Gender-specific symptoms

Similarly, participants were then asked to report whether they had experienced any short (*M* = 18.6%) or long-term (*M* = 15.2%) ‘*anomalous menstrual cycle’*. This single item measure was used to test our hypothesis.

#### Health-related covariates

To rule out potential confounds, we decided to run robustness checks including health factors that may affect menstrual cycles. First, we decided to include a measure of participants’ general health, as measured by a sum score of their self-reported medical antecedents. These were asked as a series of yes/no questions probing allergies (Oertelt-Prigione, 2012), heart (Canoy et al., 2015) and respiratory (Macsali et al., 2013) conditions, sight, digestive or neurological problems as well as mental health and sleep issues (Zukov et al., 2010) (*M* = 2.37, *SD* = 1.71). In addition, we assessed participants’ tobacco (*M* = 3.69, *SD* = 2.80) and alcohol (*M* = .93, *SD* = 1.08) consumption (0 = ‘*Never*’, 1 = ‘*Occasionally*’, 2 = ‘*At least once a week*’, 3 = ‘*Everyday*’). Overall 58.7% of the participants reported to consume tobacco daily and 2.8% alcohol.

#### Demographics

Finally, participants were asked to report their monthly income bracket (7 levels from 0 = ‘No income’ to 6 = ‘€2500 and above’, *Median* = 3, ‘*between the minimum wage and €1500*’) and diploma (7 levels from 0 = ‘*None*’ to 6 = ‘*PhD and above*’, *Median* = 2, ‘*highschool graduate*’). Participants characteristics according to their exposure group can be found in table 1.

**Table 1.** Participants characteristics according to their exposure group (n = 145)

|  | *No-exposure*<br>28(19.6)<sup>a</sup> | *< 5 times*<br>30(21)<sup>a</sup> | *5 to 10 times*<br>21(14.7)<sup>a</sup> | *> 10 times*<br>64(44.8)<sup>a</sup> |
| --- | --- | --- | --- | --- |
| *Symptoms* |  |  |  |  |
| Total General | 1.36(4.28)<sup>b</sup> | 7.00(5.02)<sup>b</sup> | 8.00(4.36)<sup>b</sup> | 9.44(3.80)<sup>b</sup> |
| LT General | 1(3.68)<sup>b</sup> | 2.43(3.78)<sup>b</sup> | 2.61(3.60)<sup>b</sup> | 3.20(3.60)<sup>b</sup> |
| Total MC | 1(3.60)<sup>a</sup> | 6(20)<sup>a</sup> | 3(14.30)<sup>a</sup> | 17(26.60)<sup>a</sup> |
| LT MC | 1(3.60)<sup>a</sup> | 5(16.70)<sup>a</sup> | 2(9.70)<sup>a</sup> | 14(21.90)<sup>a</sup> |
| *Health* |  |  |  |  |
| Antecedents | 2.82(1.74)<sup>b</sup> | 2.30(1.68)<sup>b</sup> | 2.52(1.44)<sup>b</sup> | 2.16(1.77)<sup>b</sup> |
| Tobacco | 1.82(1.44)<sup>b</sup> | 1.90(1.37)<sup>b</sup> | 1.71(1.45)<sup>b</sup> | 2.06(1.28)<sup>b</sup> |
| Alcohol | 0.68(0.67)<sup>b</sup> | 1.03(0.89)<sup>b</sup> | 0.76(0.70)<sup>b</sup> | 0.86(0.71)<sup>b</sup> |
| *Demographics* |  |  |  |  |
| Age | 53.40(7.50)<sup>b</sup> | 49.70(12.00)<sup>b</sup> | 49.30(9.24)<sup>b</sup> | 45.80(10.00)<sup>b</sup> |
| Income | 2.21(1.52)<sup>b</sup> | 3.13(1.89)<sup>b</sup> | 2.90(1.55)<sup>b</sup> | 2.45(1.76)<sup>b</sup> |
| Diploma | 1.29(0.98)<sup>b</sup> | 2.70(1.53)<sup>b</sup> | 2.52(1.50)<sup>b</sup> | 2.44(1.22)<sup>b</sup> |

*Notes.* <sup>a</sup> Numbers represent counts and number between brackets are percentages. <sup>b</sup> Numbers are means and numbers between brackets represent SD. MC = Menstrual Cycle, LT = Long-Term.

## 3. RESULTS

### 3.1. Preliminary analyses

Point-biserial and Pearson correlations were computed to investigate the links between our measures. Results can be seen in table 2 and highlight, as expected, substantial associations between teargas exposure and both total *r*(144) = .54, *p* < .001 and long-term symptom scores, *r*(144) = .35, *p* < .001. Furthermore - and providing support for our hypothesis - exposure to teargas was also positively associated with both total *r*(144) = .20, *p* = .018 and long-term menstrual cycle perturbations, *r*(144) = .17, *p* = .044.

**Table 2.** Correlation between all measured variables (n = 145)

|  | 1 | 2 | 3 | 4 | 5 | 6 | 7 | 8 | 9 | 10 | 11 |
| --- | --- | --- | --- | --- | --- | --- | --- | --- | --- | --- | --- |
| 1 Exposure | - |  |  |  |  |  |  |  |  |  |  |
| 2 T. Gen. | .54*** | - |  |  |  |  |  |  |  |  |  |
| 3 LT. Gen. | .35*** | .84*** | - |  |  |  |  |  |  |  |  |
| 4 T. MC | .20* | .51*** | .44*** | - |  |  |  |  |  |  |  |
| 5 LT. MC | .17* | .47*** | .48*** | .88*** | - |  |  |  |  |  |  |
| 6 Ante. | -.13 | .18* | .21* | .05 | .07 | - |  |  |  |  |  |
| 7 Tobacco | .07 | .05 | .02 | .05 | .07 | .11 | - |  |  |  |  |
| 8 Alcohol | .04 | -.05 | -.12 | -.02 | .01 | -.05 | .12 | - |  |  |  |
| 9 Age | -.28*** | -.16<sup>†</sup> | -.13 | -.14<sup>†</sup> | -.10 | .20* | -.14 | -.06 | - |  |  |
| 10 Income | -.01 | .01 | -.05 | -.11 | -.10 | -.12 | -.19* | -.12 | .17* | - |  |
| 11 Diploma | .23** | .19* | .02 | .05 | .05 | .05 | -.09 | .25*** | -.06 | .35*** | - |

*Notes.* Ante. = Antecedents T. = Total MC = Menstrual Cycle, LT = Long-Term. <sup>†</sup> p < .10, \* p < .05, \*\* p < .01, \*\*\* p < .001.

### 3.2. Main analysis

Although results from correlational analyses were encouraging, we needed to see if the association between teargas exposure and menstrual cycle perturbations held when using optimal analytical procedures (here logistic regression). As can be seen from table 3, logistic regression models confirmed the presence of a link between exposure to teargas and menstrual cycle perturbations, in general, O.R. = 1.62, 95%CI[1.10,2.52], *p* = .022, and for long term symptoms, O.R. = 1.56, 95%CI[1.03,2.53], *p* = .049 - albeit weaker. We also ran the models again adjusting for all health related covariates and demographics. Although none of the covariates were substantially linked with the outcome (all *ps* > .10), they did affect our main results, O.R. = 1.48, 95%CI[0.95,2.46], *p* = .088 for long-term, O.R. = 1.52, 95%CI[0.99,2.41], *p* = .061 for total menstrual cycle perturbations. This was most likely due to our small sample size and variability in the data, which rendered models sensitive to addition of unnecessary *df*s.

**Table 3.** Summary of logistics regression models for teargas exposure effects on menstrual cycle perturbations (M_1_ = Total symptoms; M_2_ = Long-term symptoms; N = 145).

| | $\chi^2$ | R<sup>2</sup> | Odds Ratio | 95%CI | Z | P |
| --- | --- | --- | --- | --- | --- | --- |
| $M_1$ | 5.99 | .04 | | | | .014 |
| *Intercept* |  |  | .21 | [.13,.33] | 6.66 | < .001 |
| *Exposure* |  |  | 1.62 | [1.10,2.52] | 2.29 | .022 |
| $M_2$ | 4.39 | .04 | | | | .036 |
| *Intercept* |  |  | .17 | [.10,.26] | 7.12 | < .001 |
| *Exposure* |  |  | 1.56 | [1.03,2.53] | 1.97 | .049 |

## 4. DISCUSSION

Could CS teargas exposure trigger menstrual perturbations among female protesters? This very first analysis suggests that teargas could be linked with menstrual cycle perturbations up to a few days after exposure. It is important to note that the present research is the first to attempt a thorough investigation of the teargas-menstrual perturbations hypothesis. However, this first study cannot account for processes and or mechanisms, so the issue of knowing whether this association may be due to the psychological or physiological effects of teargas, as described in the introduction, remains completely open. In fact, stress is known to be related to menstrual cycle disturbance (Huhmann, 2020; Xiao & Ferin, 1997). Protests, in which tear gas is used, are a very stressful event which can even lead to depression or posttraumatic stress disorder (Adam-Troian, Çelebi, & Mahfud, 2020; Celebi et al., 2020). Thus, it is possible that psychological effects cause menstrual disorders in protesters exposed to tear gas.

Another possibility would be that both psychological and physiological effects work together in causing observed symptoms. It is therefore important to better characterize more accurately when the female menstrual cycle is actually affected after exposure to tear gas, and to what extent these effects are related to physiological or psychological mechanisms. Of note, symptoms related to acute panic disorder include respiratory gasp and dizziness, therefore it is difficult to distinguishing them from lesser cyanide intoxications (Borden Institute, 2009) which can occur after exposure to CS teargas (Johnson-Kanapathy, 2013). Further studies should include more detailed measures of both exposure and symptoms (e.g. not only menstrual cycle anomalies in terms of duration but also painfulness, aspect, quantity…) which may help clarify the mechanisms at play.

Despite these theoretical limitations, the statistical associations detected by our models seemed robust, which is encouraging. Still, these were sensitive to the introduction of supplementary variables in the models, which we believe is due to various methodological reasons. First and foremost, our study was limited by sample size, due to data collection issues and this seriously limited our power and thus ability to detect small to medium effects such as the ones we found here (Baker et al., 2020). The ‘rare’ nature of our target population (very active Yellow Vests protesters) made it difficult to access participants. Moreover, we believe that our measures for both teargas exposure and menstrual cycle-related perturbations were limited (only four categories for the former, only one item for both). This probably explains the variability observed on sample average responses, and made it more difficult to detect our effects.

In addition to this detection problem, we would like to invite caution when interpreting the present findings. If these results are encouraging, they constitute preliminary and pioneer evidence in need of confirmatory studies. In fact, we do not claim the statistical association we found to have causal implication, especially due to the cross-sectional nature of our study. In addition, our sample is specific (Female Yellow Vests protesters in France), which means that generalizability beyond French protesters may not be guaranteed (Henrich, Heine, & Norenzayan, 2010). Finally, we cannot exclude that our results may be biased due to traditional limitations of self-report methods, although this may be truer for prevalence numbers (i.e. averages) than for associations between our constructs.

Within the boundaries of the above-mentioned limitations, we argue that this study provided first evidence to warrant further investigation of the potentially detrimental effect of teargas on women’s menstrual cycle and reproductive functions. We call for urgent epidemiological research to confirm or infirm whether these gender specific effects of teargas may occur, and to quantify them using larger samples of female protesters and more robust (i.e. longitudinal) designs.

## Data Availability

The data underlying our findings can be openly accessed using the following link https://osf.io/k3upf/?view_only=39158ca00ff84a03ad9d3b41691abbba

https://osf.io/k3upf/

## Notes

### Competing Interest Statement

The authors have declared no competing interest.

### Funding Statement

No funding to declare

### Author Declarations

The study was conducted in accordance with the 1964 Helsinki declaration and its later amendments, the French legislation on research involving human participants and the 2016 APA Ethical Principles of Psychologists and Code of Conduct. Ethic approval was obtained from Sehir University ethics board (n29-2019).

